# Mortality and Morbidity in Anterior Versus Posterior Circulation aSAH: A Systematic Review and Meta-Analysis

**DOI:** 10.64898/2026.03.31.26349908

**Authors:** Sai Sreedhar Tripurari, Raghavendra Nayak, Renuka A, Sreedharan Nair, Rajesh Nair, Abhinay M Huchche, Sonal Sekhar M, Vijayanarayana Kunikatta

## Abstract

**Background:** Aneurysmal subarachnoid hemorrhage (aSAH) is a severe form of stroke associated with higher morbidity and mortality. Posterior circulation aneurysms are considered to have worse prognosis than anterior circulation aneurysms due to anatomical location, hemorrhage severity, and treatment complexity. We aimed to determine whether aneurysm location independently influences clinical outcomes following aSAH

**Methods:** PubMed, Scopus, Embase, and Web of Science were searched from January 2000 to December 2025 for studies reporting outcomes in anterior or posterior circulation aSAH. The outcome analysis included mortality, functional recovery (modified Rankin Scale [mRS] 0–2 and 3–6 at 6 months and 1 year), hydrocephalus, delayed cerebral ischemia (DCI), and symptomatic cerebral vasospasm. Pooled proportions and subgroup comparisons were performed using random-effects meta-analysis (DerSimonian-Laird method). Publication bias was evaluated using contour-enhanced funnel plots and Egger’s test.

**Results:** Nineteen analytic entries from 18 studies (anterior: n = 1,625; posterior: n = 986; total N = 2,611) were included. Pooled mortality was 13% (95% CI: 10%–17%; I^2^ = 84.6%), with no significant difference between the anterior (14%; 95% CI: 10%–20%) and posterior (11%; 95% CI: 7%–18%) circulation subgroups (p = 0.437). Good functional outcome was 60% at 6 months (95% CI: 51%–67%) and 55% at 1 year (95% CI: 46%–64%), with no location-based differences. Hydrocephalus (35% vs 35%; p = 0.979) and DCI (17% vs 17%; p = 0.939) were comparable between subgroups. Symptomatic cerebral vasospasm was the only outcome differing significantly by location, occurring more frequently in anterior circulation aSAH (24% vs 11%; χ^2^ = 5.59; p = 0.018).

**Conclusion:** Aneurysm location does not independently determine mortality, functional recovery, hydrocephalus, or DCI following aSAH. Symptomatic cerebral vasospasm was the only location-specific outcome. Admission neurological grade (World Federation of Neurosurgical Societies [WFNS]), rather than vascular territory, appears to be the primary determinant of mortality. Aneurysm location alone should not guide prognostic decisions or limit aggressive treatment.

## INTRODUCTION

Rupture of an intracranial aneurysm leads to aneurysmal subarachnoid hemorrhage (aSAH), a severe form of stroke associated with poor outcomes. ^1^ Despite advancements in neurocritical care, the overall mortality rates for aSAH remain 25– 35%, with substantial survivors experiencing neurological disability. ^2^ Most aneurysmal ruptures occur in the anterior circulation, particularly at the anterior communicating artery (ACoA), middle cerebral artery (MCA), anterior cerebral artery (ACA), and internal carotid artery (ICA). In the posterior circulation, ruptures primarily occur at the basilar artery (BA), vertebral artery (VA), and posterior inferior cerebellar artery (PICA). ^3^ Posterior circulation aneurysms are considered to have worse prognosis when compared with anterior circulation due to their anatomical location, higher initial hemorrhage severity, and increased difficulty in treatment management. ^4,5^ However, the relationship between aneurysm location and clinical complications has not been systematically examined in a pooled quantitative framework, and limited guidance is available for treatment decisions based on vascular territories.

Given the persistent uncertainty in the literature and the absence of comprehensive pooled analysis, we performed a systematic review and meta-analysis to determine whether aneurysm location in anterior and posterior circulations may independently influence clinical outcomes following aSAH, including the mortality, functional recovery, hydrocephalus, DCI, and symptomatic cerebral vasospasm

## METHODS

### Search Strategy and Selection Criteria

This systematic review and meta-analysis followed the Preferred Reporting Items for Systematic Reviews and Meta-Analyses (PRISMA) statement guidelines ^6^ and was registered in PROSPERO (ID: CRD420261283547). Systematic search was conducted in PubMed, Scopus, Embase and Web of Science from January 2000 - December 2025. The search strategy was developed by using the terms related to aneurysmal subarachnoid hemorrhage, aneurysm-specific vascular locations, treatment procedures, mortality, functional outcomes, and secondary complications in aSAH. The complete search string is available in the supplementary material Inclusion criteria were as follows: (1) prospective or retrospective cohort, case series, or case-control studies reporting outcomes following aSAH confirmed by neuroimaging; (2) provision of extractable outcome data for at least one of the prespecified primary or secondary outcomes; (3) reporting of aneurysm location categorised as anterior circulation, posterior circulation, or both; (4) patients aged ≥ 18 years; (5) publication in English only. Exclusion criteria included: unruptured aneurysms, traumatic aSAH, single arterial territory studies without comparison data, and abstract-only publications.

### Outcomes Definitions

The primary outcome was to determine mortality and functional recovery in post aSAH patients. Aneurysm location was classified as anterior circulation (ACoA, MCA, ACA and PCOM originating from ICA) or posterior circulation (BA, VA, PICA, SCA, and PCA). Good functional outcomes were defined as modified Rankin Scale (mRS) 0–2 and Glasgow Outcome Scale (GOS) 4–5; poor functional outcomes as mRS 3–6 and GOS 1–3. Studies reporting multiple data points (3-month, 6-month and 1-year) for mRS and GOS were extracted preferentially. Secondary outcomes include hydrocephalus (acute, subacute and chronic) DCI and symptomatic cerebral vasospasm. Different treatment procedures for aSAH were categorised as surgical clipping or endovascular coiling.

### Data Extraction and Quality Assessment

Two independent reviewers (S.S and V.N) conducted a two-stage screening process: review of titles and abstracts followed by full-text assessment. Disagreements were resolved by discussion and consensus. Data were extracted using a standardized form capturing study characteristics, patient demographics, aneurysm location, treatment modality, risk factors, and all prespecified outcomes.

Risk-of-bias assessment was performed using Joanna Briggs Institute (JBI) critical appraisal tool for case series ^7^ and cohort studies, ^8^ which are classified as low risk (≥9/11 or ≥9/10), moderate risk (7–8), or high risk of bias (≤6) accordingly. (Table 1)

**Table 1.** Baseline Characteristics of Included Studies.

### Statistical Analysis

For each study, number of events and total sample size were extracted for each outcome. Proportions with 95% confidence intervals (CIs) were calculated using the logit transformation. The Random-effects meta-analysis using the DerSimonian-Laird (DL) method ^9^ accounted for between-study heterogeneity. Subgroup analyses compared anterior and posterior circulation groups, with between-group differences assessed using the χ^2^ test; a two-sided P<0.05 was considered statistically significant.

One study (Göttsche et al., 2022) reported outcomes separately for anterior (N = 510) and posterior (N = 93) circulation patients from the same institutional cohort, representing non-overlapping patient populations entered as separate analytic units, generating 19 analytic entries from 18 unique studies. Sensitivity analysis excluding this study from one subgroup confirmed that conclusions were not materially altered.

Heterogeneity was quantified using I^2^ statistics and between-study variance (τ^2^). ^10^ Publication bias was assessed using contour-enhanced funnel plots and Egger’s weighted regression test where n ≥10 studies contributed to an outcome. Trim-and-fill analysis was performed for outcomes with significant Egger’s test asymmetry.

A univariable meta-regression was performed to examine whether study-level DCI rate predicted log-odds of mortality. A multivariable random-effects meta-regression (REML estimator) was subsequently performed to simultaneously examine three predictors of mortality: (1) circulation territory (posterior vs anterior, as a binary variable); (2) proportion of poor-grade admission patients (WFNS IV–V or HH III–V); and (3) proportion of good-grade admission patients. This model directly tests whether location independently predicts mortality after adjusting for admission severity. Studies were included in the meta-regression only when all three predictors were available. All statistical analyses were performed using R version 4.3.2 with the meta and metafor packages.

### Standard Protocol Approvals, Registrations, and Patient Consents

Review board approval and informed patient consent were not required because this research used only published, de-identified data. The PRISMA 2020 checklist is submitted as supplementary material.

## RESULTS

### Study Characteristics

A total of nineteen analytic entries from 18 unique studies were included, published between 2011 to 2025 representing 2,611 patients for the primary mortality analysis and 2,316 patients for secondary complications. Nine entries reported anterior circulation data (n = 1,625) ^11–19^ and 10 entries reported posterior circulation data (n = 986) ^13,20–28^ (Table 1). One study (Göttsche et al., 2022) ^13^ contributed to both circulation subgroups from non-overlapping patient cohorts. A detailed study characteristics are summarized in (Table 2).

### Primary Outcome: Overall Pooled Mortality

Across all 19 analytic entries (N = 2,611), the pooled mortality was **13%** (95% CI: 10%–17%; I^2^ = 84.6%; τ^2^ = 0.358; p < 0.0001), with substantial heterogeneity attributable to differences in patient characteristics, treatment procedures, secondary complication profiles, and follow-up conditions (Figure 3A).

**Figure 1.**
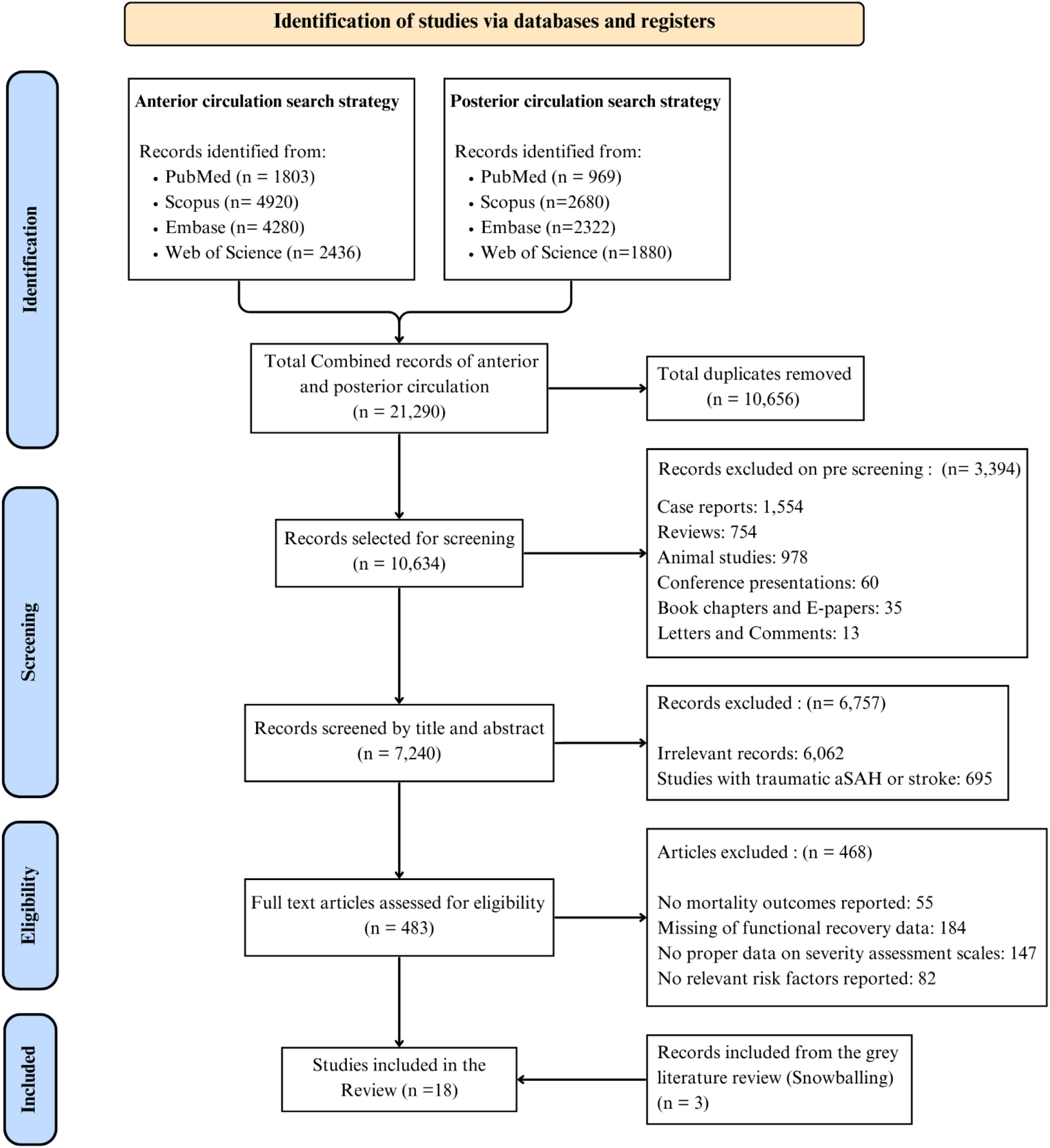
PRISMA 2020 flow diagram for systematic review of anterior and posterior circulation aSAH outcomes. A total of 18 unique studies were identified, generating 19 analytic entries; one study (Göttsche et al., 2022) contributed to both subgroups from non-overlapping patient cohorts.

**Figure 2.**
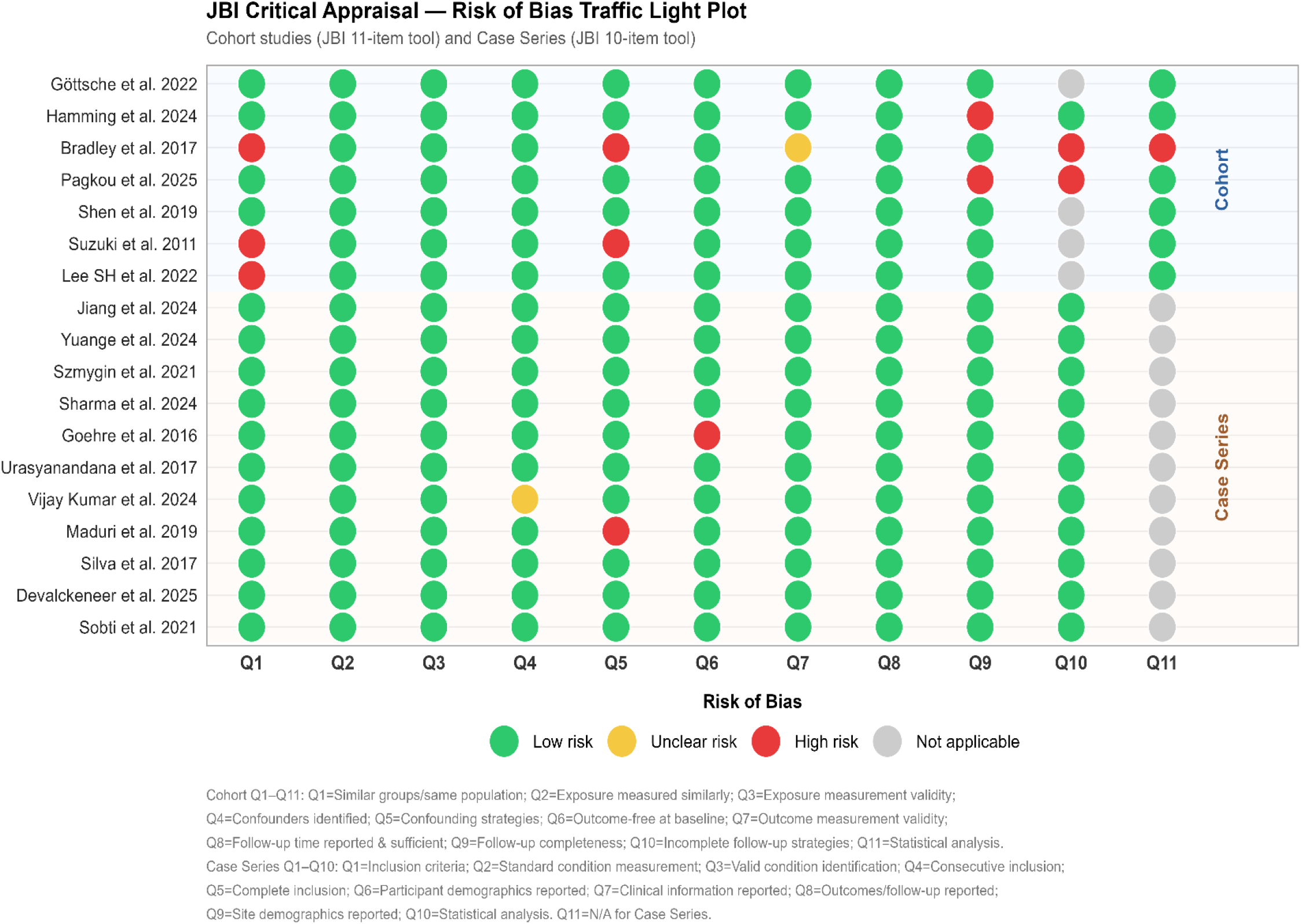
Risk of bias assessment of included studies using the JBI critical appraisal tool. Cohort studies, n = 7; case series, n = 11.

**Figure 3.**
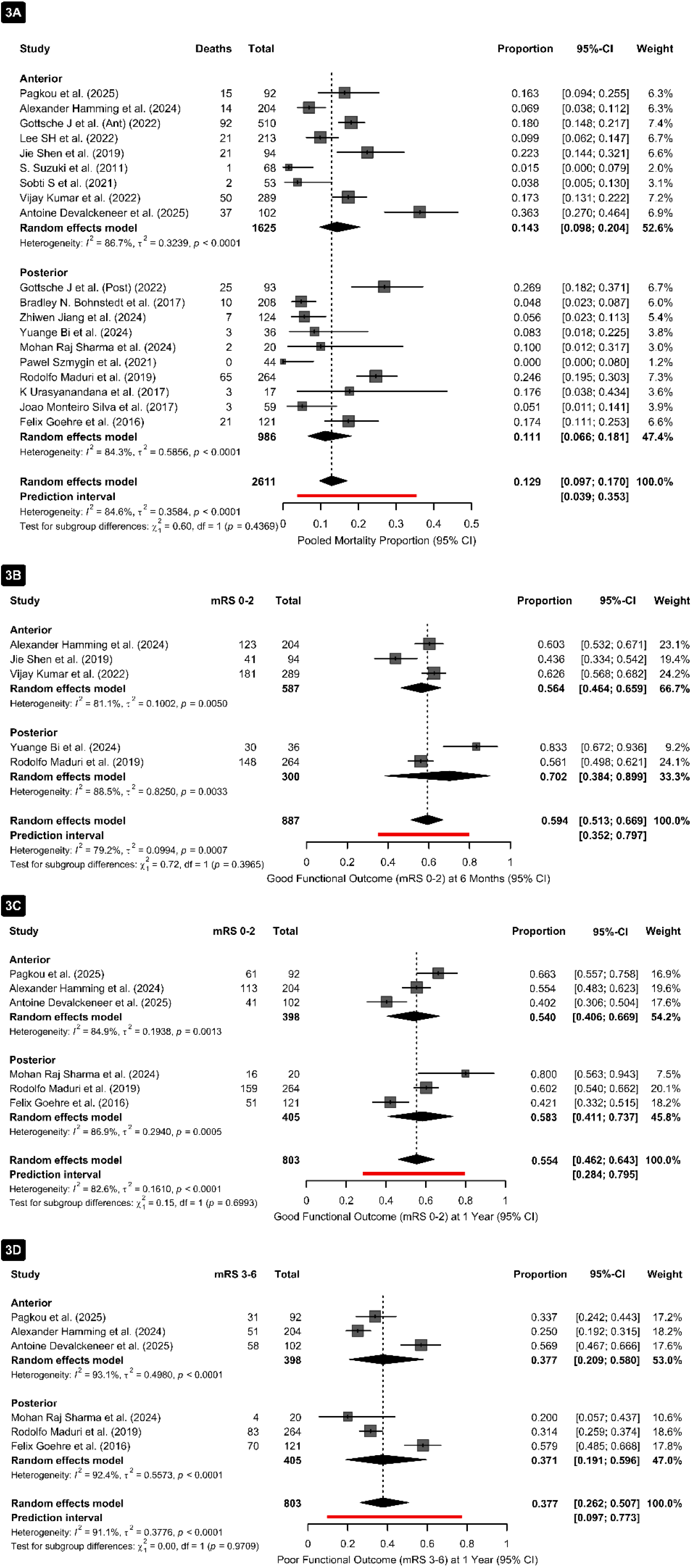
Primary clinical outcomes following aneurysmal subarachnoid hemorrhage. (3A) Pooled mortality by aneurysm location. (3B) Good functional outcome at 6 months (mRS 0–2). (3C) Good functional outcome at 1 year (mRS 0– 2). (3D) Poor functional outcome at 1 year (mRS 3–6). Squares represent study estimates; size proportional to weight. Diamonds indicate pooled random-effects estimates. Red bars indicate 95% prediction intervals. DL = DerSimonian-Laird estimator.

### Anterior Circulation Mortality

Nine analytic entries (N = 1,625) reported mortality following anterior circulation aSAH. The pooled mortality was **14%** (95% CI: 10%–20%; I^2^ = 86.7%; τ^2^ = 0.324; p < 0.0001). Individual estimates ranged from 2% (Suzuki et al., 2011; N = 68) to 36% (Devalckeneer et al., 2025; N = 102). The largest contributor, Göttsche et al. (2022; N = 510), reported mortality proportion of 18% (95% CI: 15%–22%). (Figure 3A)

### Posterior Circulation Mortality

Ten analytic entries (N = 986) reported mortality following posterior circulation aSAH. The pooled mortality was 11% (95% CI: 7%–18%; I^2^ = 84.3%; τ^2^ = 0.586; p < 0.0001). Individual estimates ranged from 0% (Szmygin et al., 2021; N = 44) to 27% (Göttsche et al. [Posterior], 2022; N = 93). (Figure 3A)

### Subgroup Comparison: Mortality

Subgroup comparison demonstrated no statistically significant difference in mortality between anterior and posterior circulation (χ^2^ = 0.60, df = 1; p = 0.437). Pooled estimates overlapped substantially, indicating that aneurysm location did not significantly influence mortality. Publication bias assessment demonstrated statistically significant funnel plot asymmetry (Egger’s test: p = 0.010), suggesting potential small-study effects (Figure 5A.1). Trim-and-fill analysis confirmed robustness of the primary estimate. (Figure 5A.2)

### Functional Recovery: Six-Month Outcomes

Five analytic entries (N = 887) reported 6-month functional outcomes. The pooled proportion achieving good functional outcome (mRS 0–2) was **60%** (95% CI: 51%– 67%; I^2^ = 79.2%) (Figure 3B). Subgroup analysis demonstrated 56% (95% CI: 46%– 66%) for anterior circulation and 70% (95% CI: 38%–90%) for posterior circulation, with no statistically significant difference (χ^2^ = 0.72; p = 0.397). (Figure 3B)

### One-Year Functional Outcomes

Six analytic entries (N = 803) reported 1-year outcomes. The overall pooled proportion for good functional outcome (mRS 0–2) was **55%** (95% CI: 46%–64%; I^2^= 82.6%; τ^2^ = 0.161) and for poor functional outcome (mRS 3–6) was **38%** (95% CI: 26%–51%; I^2^ = 91.1%; τ^2^ = 0.378) (Figures 3C–3D). Subgroup analysis showed no significant difference for good outcomes (anterior 54% vs posterior 58%; p = 0.699) or poor outcomes (anterior 38% vs posterior 38%; p = 0.971).

### Secondary Outcomes Hydrocephalus

Fifteen analytic entries (N = 2,316) reported hydrocephalus incidence. The overall pooled incidence was **35%** (95% CI: 24%–49%; I^2^ = 96.6%; τ^2^ = 1.036; p < 0.0001) (Figure 4A). Subgroup analysis demonstrated identical proportions for anterior and posterior circulation (35% vs 35%; p = 0.979), confirming that hydrocephalus risk is independent of aneurysm location. No significant publication bias was identified (Egger’s test: p = 0.189) (Figure 5B).

**Figure 4.**
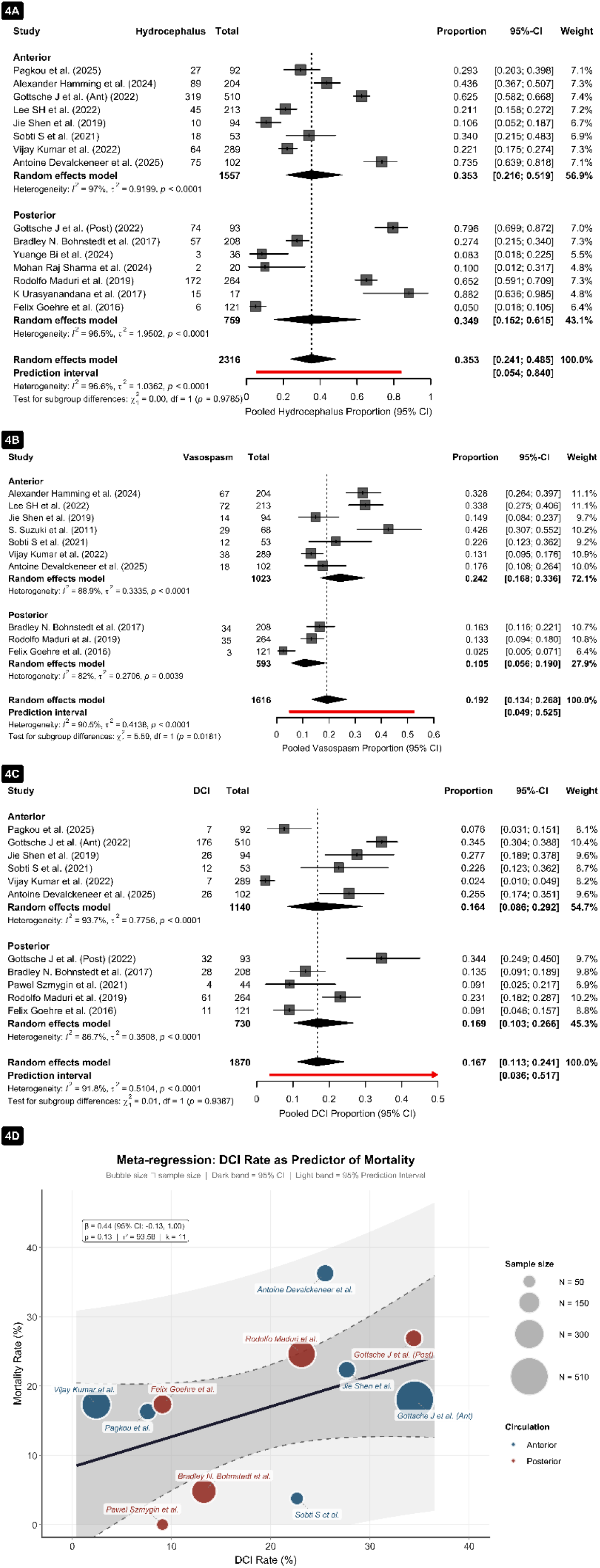
Secondary outcomes. (4A) Hydrocephalus. (4B) Cerebral vasospasm [key finding]. (4C) Delayed cerebral ischemia. (4D) Bubble plot: univariable meta-regression of study-level DCI rate as predictor of mortality. Circle size proportional to study sample size.

**Figure 5.**
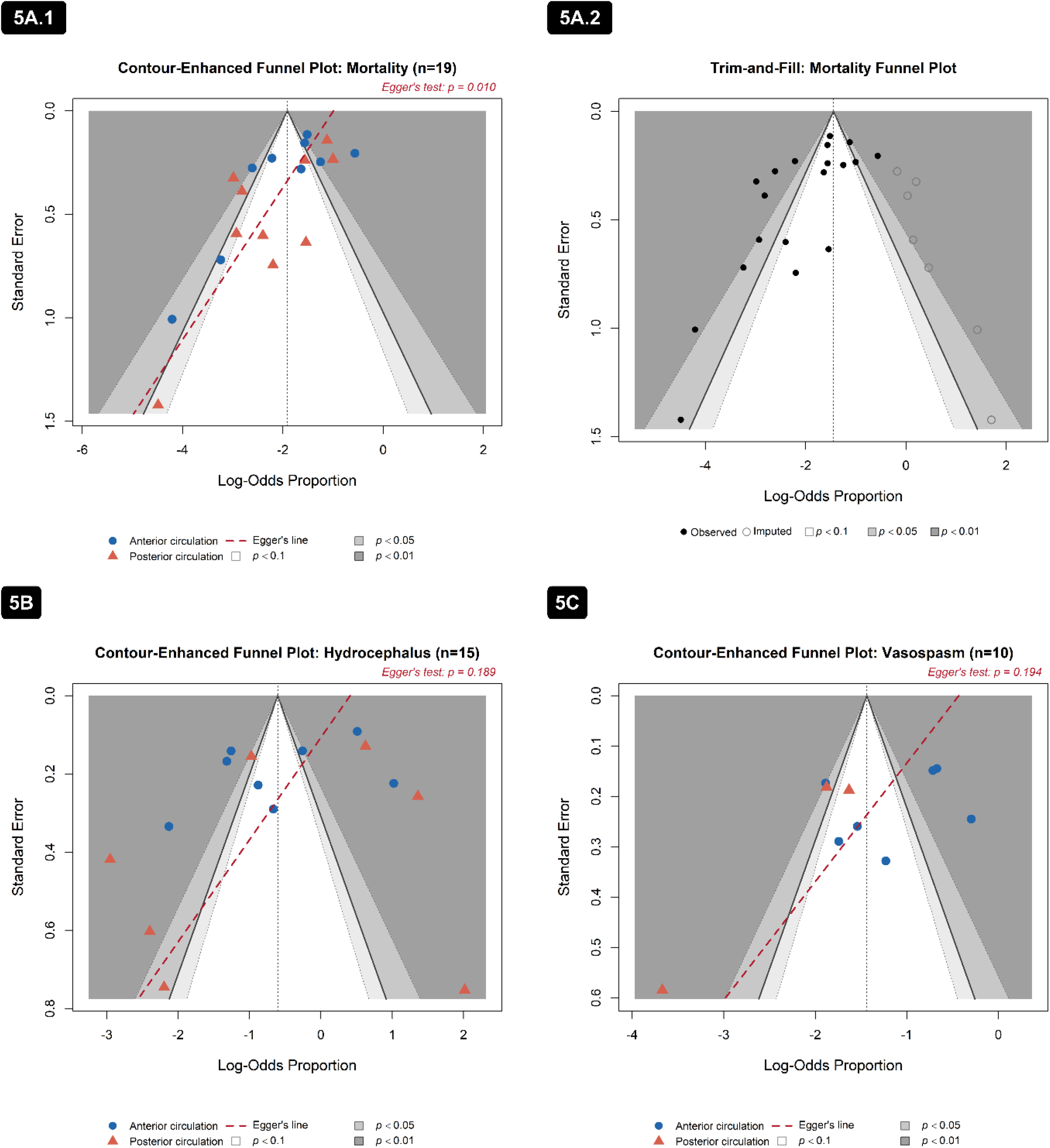
Contour-enhanced funnel plots assessing publication bias. (5A.1) Mortality (n = 19). (5A.2) *Trim-and-fill contour-enhanced funnel plot for mortality*. Filled circles = observed studies; open circles = imputed studies added by trim-and-fill analysis. (5B) Hydrocephalus (n = 15). (5C) Vasospasm (n = 10). Shaded zones: light grey = p < 0.10; medium grey = p < 0.05; dark grey = p < 0.01; white = non-significant region. Open circle = Anterior circulation; open triangle = Posterior circulation. Dashed line = Egger’s regression line.

### Symptomatic Cerebral Vasospasm

Ten analytic entries (N = 1,616) reported symptomatic cerebral vasospasm. The overall pooled proportion was **19%** (95% CI: 13%–27%; I^2^ = 90.5%) (Figure 4B). Subgroup analysis revealed significantly higher incidence in anterior circulation aneurysms (24%; 95% CI: 17%–33%) compared with posterior circulation (11%; 95% CI: 6%–19%; χ^2^ = 5.59, df = 1; p = 0.018). No significant publication bias was identified (Egger’s test: p = 0.194) (Figure 5C). The posterior vasospasm subgroup is based on n = 3 studies only and should be interpreted with caution.

### Delayed Cerebral Ischemia

Eleven analytic entries (N = 1,870) reported DCI. The overall pooled proportion was 17% (95% CI: 11%–24%; I^2^ = 91.8%; τ^2^ = 0.510) (Figure 4C). Subgroup estimates were similar (anterior 16% vs posterior 17%; χ^2^ = 0.01; p = 0.939), confirming that DCI risk does not differ by aneurysm location.

### Univariable Meta-Regression: DCI Rate as Predictor of Mortality

Univariable meta-regression examining study-level DCI rate as a predictor of log-odds of mortality demonstrated a positive but non-significant association (β = 0.44; 95% CI: −0.13 to 1.00; p = 0.13) (Figure 4D), indicating that study-level DCI rates were not consistently associated with higher mortality across included studies.

### Treatment Modality (Surgical Clipping vs Endovascular Coiling)

Subgroup analysis comparing surgical clipping and endovascular coiling demonstrated similar mortality across circulation territories. In anterior circulation, pooled mortality was **14%** for clipping and **14%** for coiling (p = 0.990). In posterior circulation, mortality was **14%** for clipping and **12%** for coiling (p = 0.683). Pooled mortality ranged from 12–14% across all location-based treatment categories, indicating no significant treatment-related mortality differences. (Figures 6A–6B).

**Figure 6.**
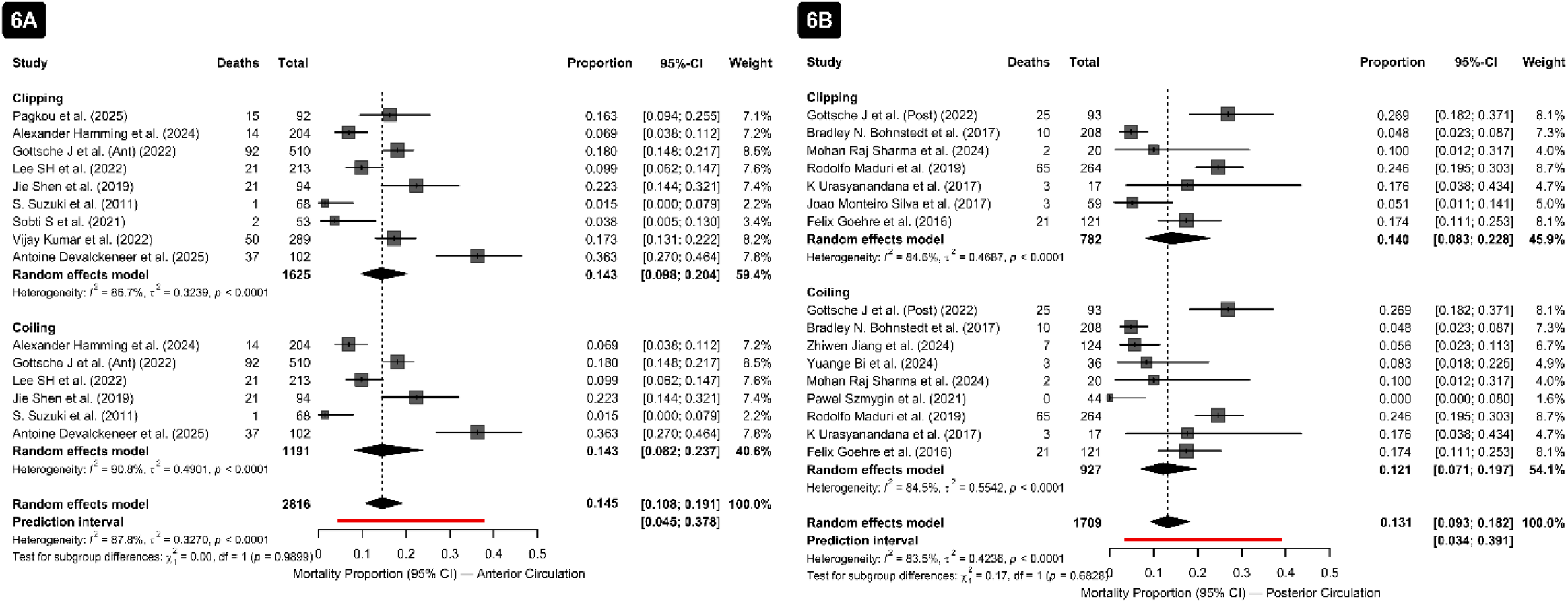
Treatment-stratified mortality analyses. (6A) Anterior circulation aneurysms stratified by treatment modality (clipping vs coiling). (6B) Posterior circulation aneurysms stratified by treatment modality. DL = DerSimonian-Laird estimator.

### Multivariable Meta-Regression: Circulation Territory, Admission Grade, and Mortality

Multivariable random-effects meta-regression was performed in studies with complete neurological grade data (n = 15), simultaneously including circulation territory (posterior vs anterior), poor-grade admission proportion (WFNS IV–V or HH III–V), and good-grade admission proportion as predictors. (Figure 7) The key finding was that circulation territory was not independently associated with log-odds of mortality after adjusting for admission grade (β = 0.105; 95% CI: −0.795 to 1.005; p = 0.819). This confirms that the previously observed numerical difference in mortality between anterior (14%) and posterior (11%) circulation subgroups is attributable to differences in patient case-mix and admission severity rather than an independent effect of vascular territory. Poor-grade admission proportion showed a clinically meaningful trend toward predicting higher mortality (β = 0.025; 95% CI:−0.003 to 0.053; p = 0.081), indicating that each percentage-point increase in the proportion of WFNS IV–V or HH III–V patients was associated with a 2.5% increase in log-odds of mortality. Good-grade admission proportion was not independently associated with mortality (β =−0.004; 95% CI: −0.026 to 0.017; p = 0.686). The model residual between-study variance was τ^2^ = 0.463. Two studies (S. Suzuki et al.; Pawel Szmygin et al.) fell outside the 95% prediction interval and were identified as potentially influential; their exclusion did not materially alter conclusions.

**7.**
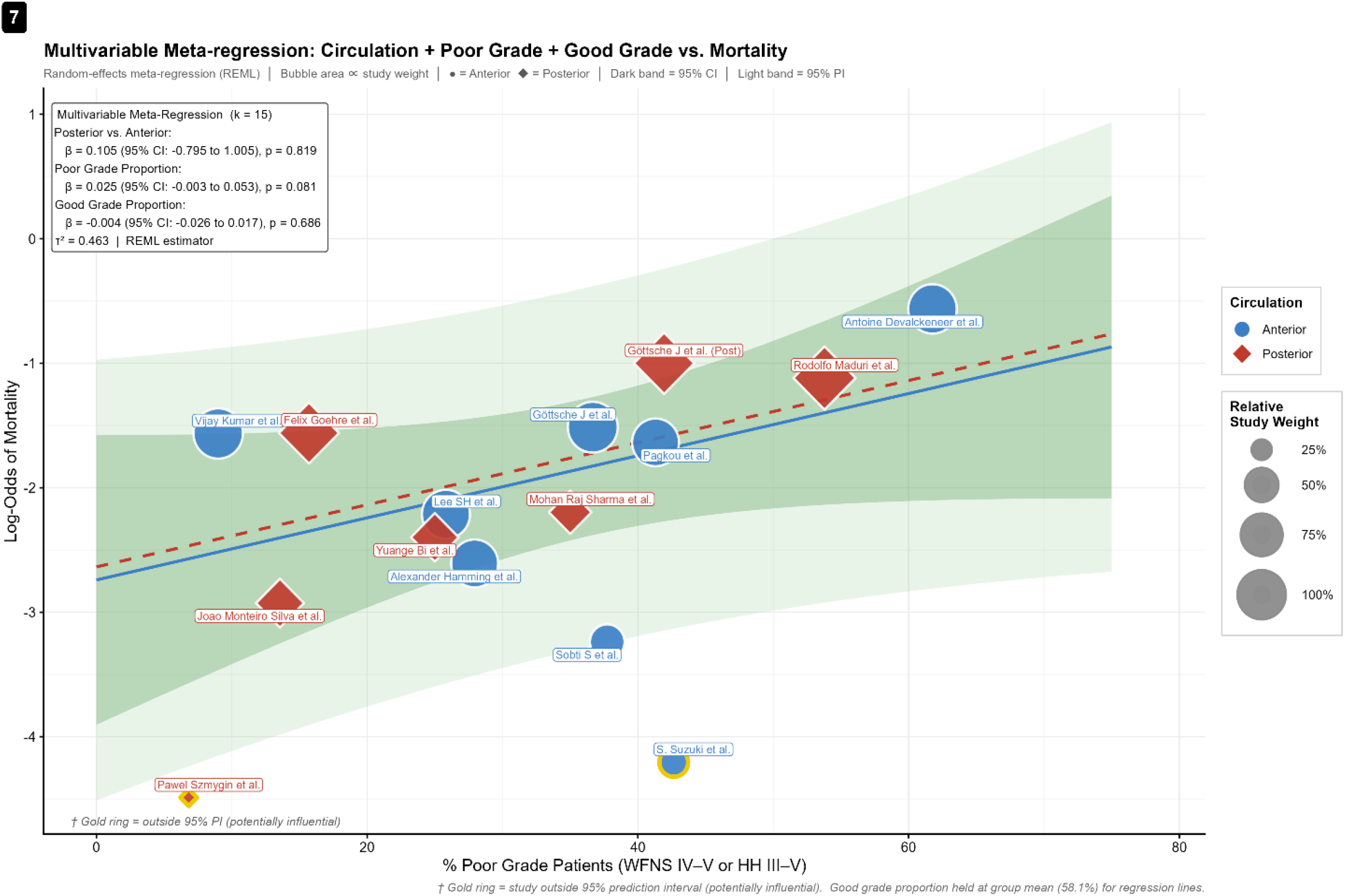
Multivariable meta-regression: circulation territory, admission grade, and mortality (k=15) X-axis: proportion of poor-grade patients (WFNS IV–V or HH III–V). Y-axis: log-odds of mortality. Blue circles = anterior circulation; red diamonds = posterior circulation. Bubble area ∝ study weight. Solid blue line = regression for anterior circulation; dashed red line = regression for posterior circulation, both at mean good-grade proportion (58.1%). Dark green band = 95% confidence interval; light green band = 95% prediction interval. Gold ring = study outside 95% PI (potentially influential). REML = restricted maximum likelihood estimator.

### Sensitivity Analysis: Leave-One-Out

Leave-one-out sensitivity analysis demonstrated that the pooled mortality estimate remained stable across all iterations. Sequential exclusion of individual studies resulted in pooled log-odds estimates ranging from −1.84 to −1.99, with all confidence intervals remaining below zero. These findings confirm that the overall mortality estimate of approximately 13% is robust and not influenced by any single study. (Figure 8)

## DISCUSSION

### Mortality: Not Independently Determined by Aneurysm Location

The pooled mortality was 13%, with estimates of 14% and 11% in anterior and posterior circulation aneurysms, respectively. This finding challenges the clinical assumption that posterior circulation aneurysms carry a substantially worse prognosis in aSAH patients. While earlier reports suggested that posterior circulation aneurysms are independently associated with higher mortality, ^29^ our pooled analysis incorporating more recent and geographically diverse studies demonstrates that the circulation-specific mortality difference is not statistically significant once studies are examined in aggregate.

Critically, multivariable meta-regression (Figure 7; n = 15) confirmed that circulation territory was not an independent predictor of mortality after adjusting for admission neurological grade (β = 0.105; p = 0.819). In contrast, poor-grade admission proportion (WFNS IV–V or HH III–V) showed a clinically meaningful trend toward predicting higher mortality (β = 0.025; p = 0.081). These findings provide direct multivariate evidence that admission neurological severity, rather than vascular territory, is the primary determinant of mortality in aSAH, consistent with the known impact of WFNS grade V which carries in-hospital mortality rates of 48–63% in large-scale studies. ^30^

### Functional Recovery: Comparable Across Both Circulation Territories

Good functional outcome (mRS 0–2) was achieved by 60% of patients at 6 months, with subgroup estimates of 56% (anterior) and 70% (posterior). At 1 year, the overall proportion was 55%, with no significant difference for either good (anterior 54%, posterior 58%; p = 0.699) or poor (anterior 38%, posterior 38%; p = 0.971) functional outcome. These findings align with population-based data reported by *Lindvall et al*., who demonstrated comparable long-term outcomes in aSAH patients independent of aneurysm location. ^31^ Prior studies reported equivalent functional recovery between circulation groups despite poor admission grades and higher rates of early hydrocephalus among posterior circulation patients, highlighting the importance of aggressive treatment regardless of aneurysm location. ^13 32^

### Hydrocephalus: A Location-Independent Complication

Hydrocephalus occurred in approximately one in three patients across both circulation groups (35% overall; anterior 35%, posterior 35%; p = 0.979), making it the most common secondary complication identified in this analysis, irrespective of aneurysm rupture location. The reported incidence of hydrocephalus after aSAH in the literature ranges from 20% to 48% depending on the phase of onset. ^33,34^ The present analysis examined pooled proportions across all time points, with extremely high heterogeneity (I^2^ = 96.6%), indicating significant differences in how hydrocephalus was defined, diagnosed, and reported across included studies. This heterogeneity reflects methodological variation (acute vs chronic hydrocephalus definitions) rather than evidence of location-specific risk.

### Delayed Cerebral Ischemia: A Shared Risk Regardless of Location

DCI occurred in 17% of patients with no significant difference between anterior (16%) and posterior (17%) circulation groups (p = 0.939), indicating that approximately 1 in 6 aSAH patients developed DCI. This aligns with existing literature, where DCI is reported in approximately 16–40% of aSAH patients depending on diagnostic criteria. ^35^ The meta-regression examining DCI rate as a mortality predictor showed a positive but non-significant association (β = 0.44; 95% CI: −0.13 to 1.00; p = 0.13) which may reflect confounding by clinical severity. These findings support previous work suggesting that DCI is one of the most important determinants of unfavorable neurological outcome after aSAH. ^36^ The substantial residual heterogeneity (I^2^ = 91.8%; τ^2^ = 0.5104) likely reflects differences in DCI diagnostic criteria and timing of assessment across studies.

### Cerebral Vasospasm: The One Location-Specific Finding

Symptomatic cerebral vasospasm was the only secondary outcome demonstrating a statistically significant difference between anterior and posterior circulation aSAH (24% vs 11%; χ^2^ = 5.59; p = 0.018), with no evidence of publication bias (Egger’s test: p = 0.194). This finding indicates that anterior circulation aSAH confers approximately twice the risk of symptomatic vasospasm compared with posterior circulation aSAH, attributable to greater exposure of proximal vessels to subarachnoid blood and vasospastic mediators within the basal cisterns. ^4^ A recent systematic review and meta-analysis of over 19,000 patients reported that when imaging was performed on clinical indication, approximately one in three patients with aneurysmal subarachnoid hemorrhage developed vasospasm. ^37^ While patients with posterior circulation aSAH are not exempt from vasospasm risk, they represent a lower-risk subgroup. The posterior vasospasm subgroup is based only on 3 studies (N = 393 patients), carrying substantial uncertainty. These finding warrants confirmation in larger prospective studies with standardized vasospasm detection criteria.

### Treatment Modality: No Location-Specific Mortality Differential

Neither surgical clipping nor endovascular coiling demonstrated a statistically significant mortality benefit across both the circulation territory. Pooled mortality of 12–14% across location-based treatment categories indicate similar outcomes, likely reflecting advancements in both surgical and endovascular techniques from 2011 to 2025. This finding aligns with previous meta-analyses demonstrating comparable mortality between clipping and coiling despite potential differences in functional outcomes. ^38,39^

The leave-one-out sensitivity analysis confirmed that the pooled mortality estimate of approximately 13% was consistent and stable across all iterations, with log-odds confidence intervals remaining entirely below zero, supporting the reliability of the overall estimate.

### Clinical and Methodological Implications of the Multivariable Meta-Regression

The multivariable meta-regression represents the key methodological advance of this study beyond prior systematic reviews. By simultaneously adjusting for circulation territory and admission neurological grade, this analysis directly addresses the fundamental confounding problem in aSAH research: posterior circulation aneurysms may appear to have worse outcomes partly because they more often present with higher clinical grades. Our model demonstrates that after accounting for this confounding, the location effect on mortality is negligible (β = 0.105, p = 0.819), while poor admission grade carries a clinically meaningful statistically marginal mortality signal (β = 0.025, p = 0.081). This finding directly supports the clinical recommendation that aggressive treatment should not be withheld based on aneurysm location alone.

### Why High Heterogeneity Does Not Preclude Pooling

The substantial heterogeneity observed across multiple outcomes (I^2^ = 84.6–96.6%) warrants explicit justification for pooled estimation. Pooling remains appropriate and clinically meaningful for the following reasons: (1) the research question concerns the direction and magnitude of location-based difference, not a precise point estimate; (2) random-effects models appropriately accommodate between-study variance; (3) 95% prediction intervals (shown as red bars in forest plots) provide conservative estimates of the plausible range across future studies; (4) sources of heterogeneity (baseline severity, treatment protocols, outcome definitions, study design) are clinically documented and expected; and (5) the multivariable meta-regression accounts for key sources of heterogeneity including admission grade. ^10^

### Study Novelty and Contribution

To our knowledge, this is the first comprehensive systematic review and meta-analysis to assess the impact of aneurysm location on a broad range of aSAH outcomes including mortality, functional recovery at 6 and 12 months, hydrocephalus, DCI, vasospasm, and treatment-related mortality in a single pooled quantitative framework using contemporary systematic review methodology. The use of contour-enhanced funnel plots, Egger’s tests, trim-and-fill analysis, meta-regression, and leave-one-out sensitivity analysis provides a robust framework for assessing potential biases and exploring outcome predictors.

## LIMITATIONS

Several limitations should be considered when interpreting these findings.

First, aneurysm locations are dichotomized into anterior and posterior circulation groups. While enabling robust statistical analysis, this may not fully capture clinical differences between individual aneurysm sites regarding hemorrhage distribution, vasospasm susceptibility, and treatment complexity, which may independently influence outcomes. Future studies with individual aneurysm site data (ACoA vs MCA vs PICA vs BA) would enable more granular analysis.

Second, the majority of included studies were retrospective observational analyses susceptible to selection bias and residual confounding. When adjusted estimates were unavailable, crude estimates were pooled with adjusted ones.

Third, substantial heterogeneity was observed across several outcomes (I^2^ = 84.6– 96.6%). Random-effects models and prediction intervals account for this variability, but important clinical moderators such as Modified Fisher scale, aneurysm morphology, and rupture-to-treatment interval could not be fully examined due to inconsistent reporting. Egger’s test demonstrated significant funnel plot asymmetry for mortality (p < 0.010); however, trim-and-fill analysis did not materially alter the pooled estimate.

Fourth, functional outcome data were limited. mRS-based outcomes were available in only 5 studies at 6 months and 6 studies at 1 year, reducing statistical reliability.

Fifth, one study (Göttsche et al., 2022)^10^ contributed data to both circulation subgroups, introducing non-independence in the between-subgroup chi-squared test. Sensitivity analyses confirmed robustness of conclusions.

Sixth, the multivariable meta-regression was limited to n = 15 studies with complete grade data, and two studies were identified as potentially influential (outside the 95% prediction interval). The REML estimator was used for meta-regression, which is standard practice but differs from the DL estimator used for the proportion meta-analyses. These limitations should be considered when interpreting the meta-regression findings.

## CONCLUSIONS

In patients with aneurysmal subarachnoid hemorrhage, aneurysm location did not independently determine mortality, functional recovery, hydrocephalus, or delayed cerebral ischemia. Pooled mortality was consistent at approximately 13% across all location and treatment subgroups, with stable results in leave-one-out sensitivity analysis, regardless of whether patients were treated with surgical clipping or endovascular coiling.

These findings do not support the use of aneurysm location alone to guide prognostic decisions or to limit aggressive treatment, particularly in posterior circulation aSAH. Hemorrhage severity at presentation (WFNS grade), rather than vascular territory, appears to be the primary determinant of outcome.

Cerebral vasospasm was the single outcome that differed significantly by location, occurring at nearly twice the rate in anterior compared with posterior circulation aSAH (24% vs 11%; p = 0.018). However, this difference was statistically significant, it should not preclude aggressive monitoring and management of vasospasm in posterior circulation aSAH.

Overall, hemorrhage severity and secondary complication management appear to be the primary determinants of outcome in aSAH rather than aneurysm location. Prospective multicenter studies with standardized outcome definitions, individual aneurysm site data, and consistent reporting of baseline severity scales are needed to further characterize the anatomical factors influencing prognosis in aSAH.

## Data Availability

The data used in this systematic review and meta-analysis were extracted from previously published studies, all of which are publicly available in the cited references. The complete search strategy is provided in the Supplementary Material. The dataset of extracted study-level data and the R code used for all statistical analyses are available from the corresponding author (V. Kunikatta) upon reasonable request.

## Sources of Funding

This research received no specific grant from any funding agency in the public, commercial, or not-for-profit sectors.

## Disclosures

S.S. Tripurari, R. Nayak, Renuka A, S. Nair, R.P. Nair, A.M. Huchche, M.S. Sekhar, and V. Kunikatta report no disclosures.

## Notes

### Competing Interest Statement

The authors have declared no competing interest.

### Clinical Trial

NA

### Funding Statement

This research received no specific grant from any funding agency in the public, commercial, or not-for-profit sectors. The authors declare that no payment or services were received from any third party for any aspect of this work, including study design, data collection, statistical analysis, or manuscript preparation.

